# Cognitive and Neuroimaging Biomarker Intra-Individual Variability in Alzheimer’s Disease

**DOI:** 10.64898/2026.06.21.26356195

**Authors:** Min Huey Teo, Malcolm Ren Qing Taong, Cheuk Ni Kan, Chin Hong Tan, the Alzheimer’s Disease Neuroimaging Initiative

## Abstract

**Background:** Greater cognitive intra-individual variability (IIV) reflects increased heterogeneous performance across cognitive domains and has been linked to a higher risk of Alzheimer’s disease (AD). However, it remains unclear whether cognitive IIV is linked to heterogeneous dispersion of regional AD pathology. Hence, we aimed to examine the association between cognitive IIV and AD neuroimaging biomarker IIV.

**Methods:** This study included participants with normal cognition (CN) and mild cognitive impairment (MCI) from the Alzheimer’s Disease Neuroimaging Initiative. Cognitive IIV was computed as the within-person standard deviation of five domain-specific neuropsychological test z-scores. Four neuroimaging biomarker IIV metrics were similarly derived using regional amyloid-β (n = 1,021), tau (n = 719), cortical thickness (n = 2,148), and combined amyloid-tau-neurodegeneration (ATN, n = 258). Associations between cognitive IIV and each biomarker IIV were evaluated using linear regression models, adjusted for relevant covariates.

**Results:** Higher cognitive IIV was associated with greater biomarker IIV across amyloid-β (β = 0.039, *SE* = 0.014, *p* = .006), tau (β = 0.196, *SE* = 0.033, *p* < .001), cortical thinning (β = 0.036, *SE* = 0.008, *p* < .001), and ATN (β = 0.176, *SE* = 0.043, *p* < .001). Interaction analyses revealed that the associations of cognitive IIV with tau IIV, cortical thickness IIV, and ATN IIV were stronger in MCI than CN individuals. Significant interactions between cognitive IIV and biomarker positivity status showed that the effect with amyloid-β IIV was attenuated in A- (β = 0.004, *SE* = 0.014, *p* = .78) but that the effect with tau IIV remained robust even in T-individuals (β = 0.088, *SE* = 0.022, *p* < .001).

**Conclusion:** Elevated cognitive IIV is associated with greater heterogeneity in cortical dispersion of AD-related pathology, particularly in prodromal AD and in the presence of abnormal pathology. As a novel measure that captures variation in topographical scattering of AD pathological burden across the cortex, AD biomarker IIV may offer research and clinical utility beyond evaluating absolute biomarker load or thresholds.

## Introduction

Alzheimer’s disease (AD) is a neurodegenerative disease characterized by the accumulation of amyloid-β plaques, neurofibrillary tangles, synaptic dysfunction, and neuronal loss culminating in medial temporal to neocortical atrophy [1]. The neuropathological features may begin decades before overt clinical deficits that progressively result in the dementia syndrome. Just as the distribution of neurofibrillary tangles follows a regionally specific pattern in the staging of AD [2], its cognitive manifestation typically begins with an early decline in episodic memory, visuospatial processing, and verbal fluency [3, 4]. However, these group-level trajectories of cognitive dysfunction may fail to capture the substantial variation in domain-specific cognitive decline across different individuals. Consequently, increasing attention has been directed toward alternative approaches to operationalizing traditional cognitive measures for the early identification of AD.

Cognitive intra-individual variability (IIV), defined as the dispersion of performance across cognitive tests spanning multiple domains measured at a single timepoint, reflects the degree of variability in an individual’s cognitive profile that may be indicative of early dysfunction [5]. Elevated cognitive IIV has been associated with mild cognitive impairment (MCI) and AD genetic risk [6], and has been shown to be associated with negative outcomes such as cognitive and functional decline, progression to MCI, and incident AD [7–18]. The predictive effects of cognitive IIV were also found to be comparable to the effects of established cerebrospinal fluid (CSF) biomarkers of tau and amyloid-β [9], highlighting its close relevance to AD vulnerability and its potential as a more sensitive neuropsychological marker than conventional measures of mean cognitive performance.

While the neural pathways underlying cognitive IIV are not well understood, elevated cognitive IIV has been associated with neuropathological burden of neurofibrillary tangles [19] and neuroimaging biomarkers such as increased medial temporal lobe atrophy, amyloid- β, tau, and glucose metabolism [20–22]. Although evidence suggests that these associations between neuroimaging abnormalities and elevated cognitive IIV are strongest in frontotemporal regions [20], consistent with the stage-dependent spread of AD pathology [2], it remains unclear whether greater cognitive IIV corresponds to increased spatial dispersion of cerebral pathology across multiple brain regions. Elucidating this association may provide important insights into the utility of quantifying AD biomarker IIV beyond absolute pathological load or categorical staging in AD.

Consequently, we aimed to investigate the association between cognitive IIV and AD neuroimaging biomarker IIV across regional amyloid-β (A), tau (T), neurodegeneration (N), and combined ATN measures. We hypothesized that higher cognitive IIV would be associated with greater neuroimaging IIV across all biomarkers, with stronger effects in individuals with greater clinical severity and pathological burden.

## Methods

### Participants

Data for this study were drawn from the Alzheimer’s Disease Neuroimaging Initiative (ADNI) database (adni.loni.usc.edu). The ADNI was launched in 2003 as a public-private partnership, led by Principal Investigator Michael W. Weiner, MD. The primary goal of ADNI has been to test whether serial magnetic resonance imaging (MRI), positron emission tomography (PET), other biological markers, and clinical and neuropsychological assessment can be combined to measure the progression of mild cognitive impairment (MCI) and early Alzheimer’s disease (AD).

We selected baseline data from participants who were diagnosed as cognitively normal (CN) and MCI across all ADNI phases. Baseline clinical diagnosis in ADNI was determined based on clinician judgment informed by established clinical assessments, including the Clinical Dementia Rating (CDR), Mini-Mental State Examination (MMSE), and Wechsler Logical Memory II. Participants diagnosed with dementia and those with missing neuropsychological scores necessary to compute cognitive IIV and covariates (i.e., age, sex, education, or diagnosis) were excluded. The resulting sample sizes were comparable to or larger than those used in prior neuroimaging studies examining cognitive IIV and AD-related biomarkers [20].

To maximize statistical power, we conducted separate analyses based on data availability for each biomarker: Amyloid-β analysis included 1,021 participants (508 CN, 513 MCI) from ADNIGO, ADNI2, ADNI3, and ADNI4 with florbetapir PET scans; Tau analysis included 719 participants (459 CN, 260 MCI) from ADNI3 and ADNI4 with flortaucipir PET scans; Cortical thickness analysis included 2,148 participants (986 CN, 1,162 MCI) from all ADNI phases with usable structural MRI scans. A subset of 258 participants (178 CN, 80 MCI) from ADNI3 and ADNI4 who had complete amyloid-β, tau, and cortical thickness data were included in the ATN analyses. Participants with incomplete biomarker data required for a given analysis were excluded from that specific analysis. Key demographics of each biomarker sample are summarized in Table 1.

**TABLE 1.** Key demographics of the 4 biomarker IIV samples.

| | Aβ PET | | Tau PET | | Cortical Thickness | | ATN | |
| --- | --- | --- | --- | --- | --- | --- | --- | --- |
|  | CN (*n*=508) | MCI (*n*=513) | CN (*n*=459) | MCI (*n*=260) | CN (*n*=986) | MCI (*n*=1162) | CN (*n*=178) | MCI (*n*=80) |
| Age, M (SD) | 70.98 (6.92) | 71.77 (7.51) | 69.06 (6.90) | 72.16 (7.54) | 71.65 (6.91) | 72.65 (7.66) | 68.95 (7.29) | 72.68 (7.47) |
| Education, M (SD) | 16.42 (2.49) | 16.30 (2.61) | 16.57 (2.31) | 16.26 (2.43) | 16.45 (2.55) | 15.93 (2.76) | 16.42 (2.37) | 16.49 (2.47) |
| Sex, female (%) | 314 (61.81) | 241 (46.98) | 304 (66.23) | 123 (47.31) | 586 (59.43) | 498 (42.86) | 125 (70.22) | 37 (46.25) |
| Cognitive IIV, M (SD) | 0.70 (0.32) | 0.78 (0.41) | 0.70 (0.33) | 0.83 (0.44) | 0.69 (0.33) | 0.79 (0.40) | 0.68 (0.31) | 0.83 (0.53) |
| Amyloid-β IIV, M (SD) | 0.35 (0.16) | 0.41 (0.18) | - | - | - | - | - | - |
| Tau IIV, M (SD) | - | - | 0.39 (0.19) | 0.64 (0.50) | - | - | - | - |
| Cortical Thickness IIV, M (SD) | - | - | - | - | 0.73 (0.13) | 0.79 (0.15) | - | - |
| ATN IIV, M (SD) | - | - | - | - | - | - | 0.70 (0.17) | 0.97 (0.43) |

Abbreviations: ATN = Amyloid-Tau-Neurodegeneration, CN = cognitively normal, MCI = mild cognitive impairment.

### Cognitive intra-individual variability

Cognitive IIV was derived using multi-domain neuropsychological tests, including the Trail Making Test (TMT) A (processing speed) and B (executive function), the Rey Auditory Verbal Learning Test (RAVLT) Immediate Recall and Forgetting (memory), and Animal Fluency (language). These measures were available across all ADNI phases and used in a prior study [20]. For TMT-A, TMT-B, and RAVLT Forgetting, scores were reversed such that lower scores consistently reflected poorer cognitive performance. Only participants with complete data across all five cognitive measures were included. Raw scores of each cognitive test were standardized separately based on the distribution of each biomarker sample and cognitive IIV of each participant was computed as the standard deviation of the five test-specific z-scores at baseline. This computation is consistent with prior work operationalizing within-person variability across neuropsychological measures [6, 11] and has better statistical properties compared to the highly correlated coefficient of variation (CoV) measure [5]. Higher cognitive IIV values reflect greater heterogeneity in cognitive performance across domains.

### Neuroimaging processing and biomarker intra-individual variability

Cerebral amyloid-β and tau were assessed using florbetapir (AV-45) and flortaucipir (AV-1451) PET imaging, respectively. Neurodegeneration was assessed using cortical thickness derived from structural MRI. Details of the image processing procedure and quality control have been extensively documented [23–25]. Briefly, cortical reconstruction was performed using FreeSurfer and regional cortical thickness was delineated based on the Desikan-Killiany atlas [26]. The PET images were co-registered to each participant’s structural MRI and standardized uptake value ratio (SUVR) values were calculated across the 68 delineated bilateral cortical regions, normalized against the whole cerebellum and inferior cerebellum for amyloid-β SUVR and tau SUVR respectively. Similar to cognitive IIV, biomarker IIV was then computed within person as the standard deviation of these regional z-scores, deriving separate amyloid-β, tau, and cortical thickness IIV measures. For a subset of participants with complete amyloid-β, tau, and cortical thickness data, ATN IIV was computed using a similar approach utilizing all 204 regional z-scores (68 amyloid-β, 68 tau, and 68 cortical thickness regions). For each biomarker IIV, individual regional values were standardized based on the distribution of the respective sample. Figure 1 illustrates the differences in regional tau SUVR between two demographically-matched MCI participants with high and low tau IIV.

**Fig. 1.**
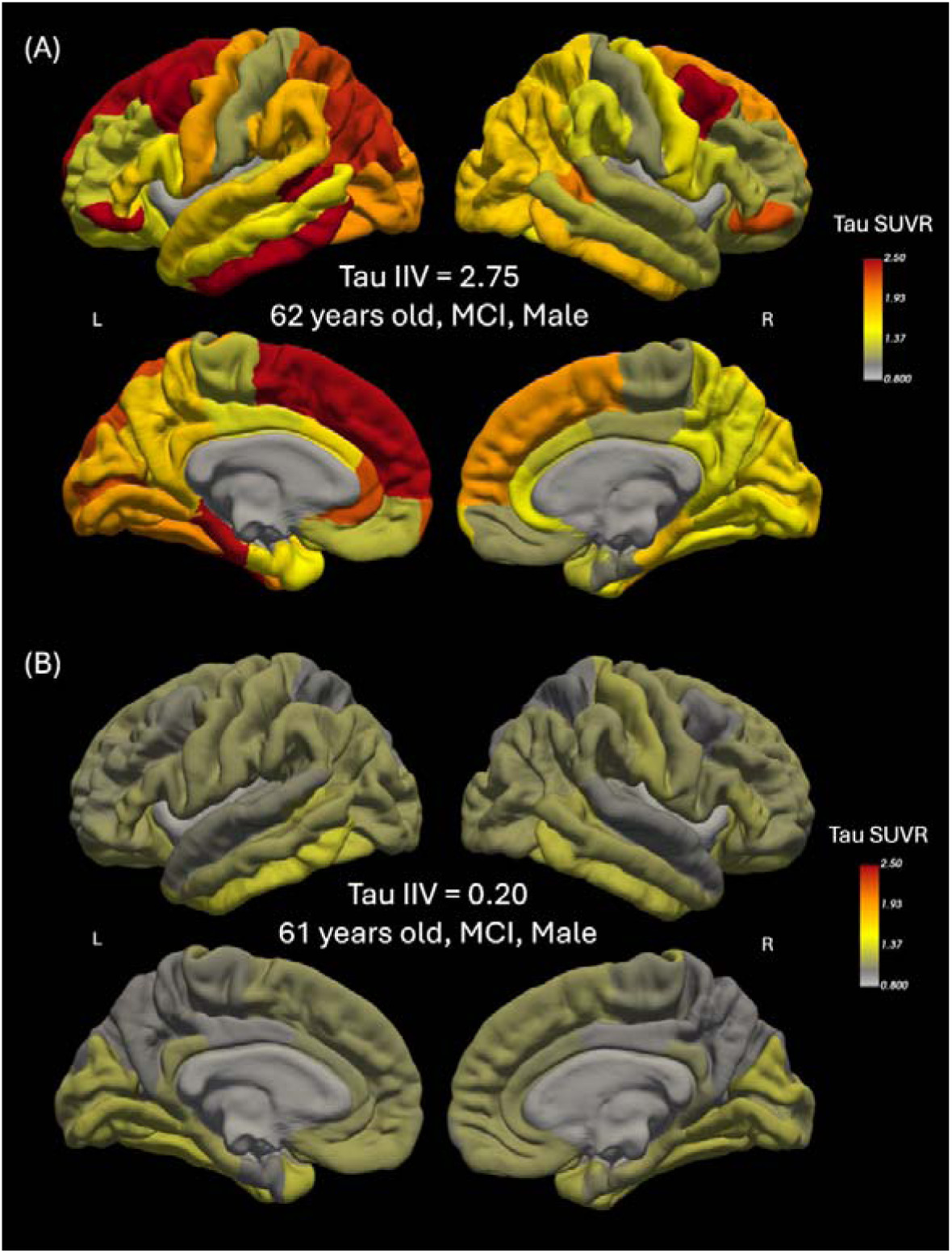
Representative illustration of regional heterogeneity in tau SUVR. Between two demographically-matched MCI participants, high tau IIV **(A)** showed pronounced regional heterogeneity in tau uptake, whereas low tau IIV **(B)** showed comparatively less heterogeneous cortical tau distribution.

### Statistical analysis

First, we conducted multiple linear regression to test whether cognitive IIV was associated with biomarker IIV in amyloid-β, tau, cortical thickness, and ATN in separate models. All models were adjusted for age, sex, years of education, and diagnosis (CN/MCI). To examine whether associations differed by clinical severity for each biomarker, we then included interaction terms between cognitive IIV and clinical diagnosis (CN/MCI). Lastly, we conducted additional interaction analyses to test whether the associations between cognitive IIV and biomarker IIV differed by amyloid-β (A-/A+) and tau (T-/T+) status, controlling for the same covariates. A+ was defined as having a global amyloid-β SUVR of more than 1.11 [27] and T+ was defined as having a meta-temporal tau SUVR of more than 1.80 [28]. For statistically significant interactions, we followed up with simple slopes analyses to decompose the interaction effects. Finally, to account for multiple comparisons, we applied false discovery rate (FDR) correction separately to the main effects, diagnosis interaction models, and biomarker-status interaction models. All reported p-values are uncorrected; however, the pattern of statistical significance remained unchanged after FDR correction. All statistical analyses were conducted in R (Version 4.5.1).

## Results

### Association of cognitive IIV with amyloid-β IIV

Higher cognitive IIV was associated with higher amyloid-β IIV, reflecting greater heterogeneous dispersion of amyloid-β across cortical regions (β = 0.039, *SE* = 0.014, 95% CI = [0.011, 0.066], *p* = .006). Interaction analyses showed that this association was not statistically influenced by clinical severity (β*_INT_* = 0.039, *SE* = 0.029, 95% CI = [-0.018, 0.095], *p* = .178; Figure 2A) but was influenced by amyloid-β positivity status (β*_INT_* = 0.052, *SE* = 0.022, 95% CI = [0.009, 0.094], *p* = .017; Figure 3A). Simple slopes analysis showed that higher cognitive IIV was associated with higher amyloid-β IIV only in A+ (β = 0.055, *SE* = 0.016, 95% CI = [0.023, 0.088], *p* < .001) but not A- (β = 0.004, *SE* = 0.014, 95% CI = [- 0.024, 0.032], *p* = .78).

**Fig. 2.**
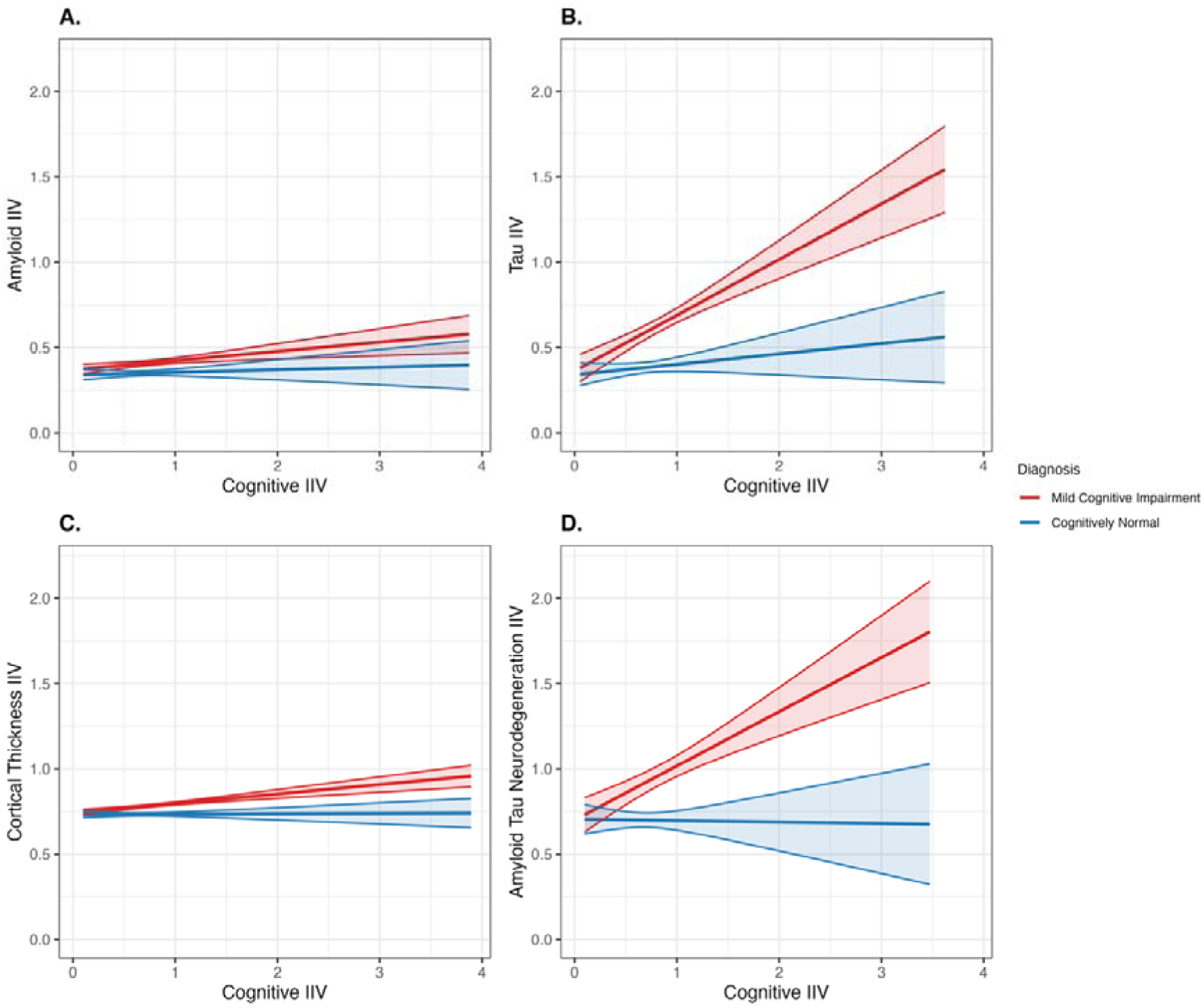
Interactive effect of clinical severity on the association of cognitive IIV and AD neuroimaging biomarker IIV. Effects between higher cognitive intraindividual variability (IIV) and AD biomarker IIV were amplified in MCI individuals for tau **(B)**, cortical thickness **(C)**, and ATN (**D)**, but not amyloid-β **(A)**, when compared to cognitively normal (CN) individuals.

**Fig. 3.**
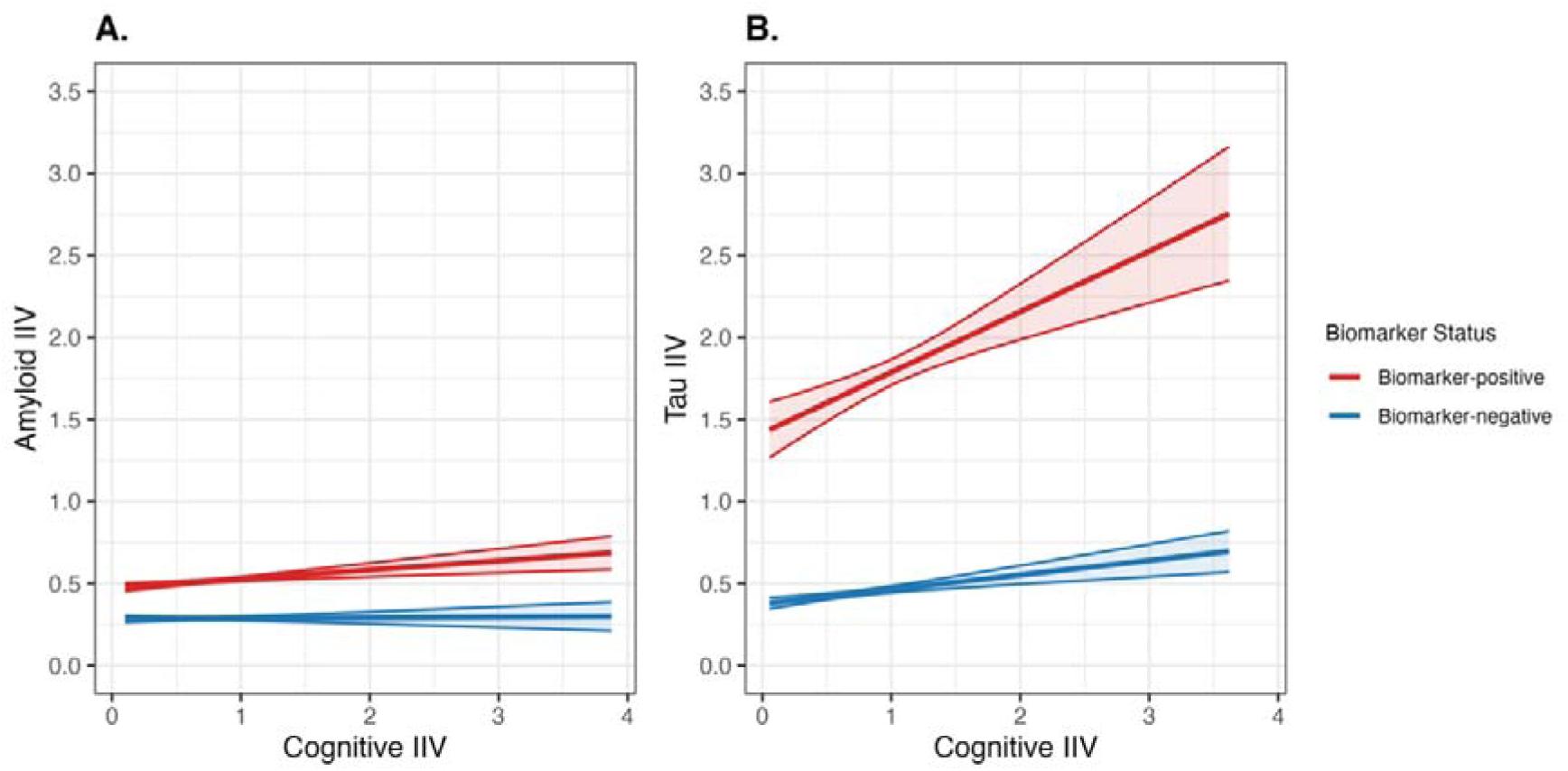
Interactive effects of biomarker status on the associations of cognitive IIV with neuroimaging IIV. Effects between higher cognitive intraindividual variability (IIV) and biomarker IIV were stronger in amyloid-β positive (**A)** and tau positive (**B)** individuals.

### Association of cognitive IIV with tau IIV

Higher cognitive IIV was associated with higher tau IIV, indicating greater heterogeneity in tau dispersion across cortical regions (β = 0.196, *SE* = 0.033, 95% CI = [0.131, 0.261], *p* < .001). There was a statistically significant interaction with clinical severity (β*_INT_* = 0.265, *SE* = 0.065, 95% CI = [0.138, 0.393], *p* < .001; Figure 2B), where higher cognitive IIV was associated with higher tau IIV in MCI (β = 0.326, *SE* = 0.046, 95% CI = [0.237, 0.416], *p* < .001) but not in CN individuals (β = 0.061, *SE* = 0.046, 95% CI = [-0.030, 0.152], *p* = .19). The association was also influenced by tau positivity status (β*_INT_* = 0.281, *SE* = 0.082, 95% CI = [0.121, 0.441], *p* < .001; Figure 3B), where the effect between higher cognitive IIV and tau IIV was stronger in T+ (β = 0.369, *SE* = 0.079, 95% CI = [0.215, 0.524], *p* < .001) than in T- (β = 0.088, *SE* = 0.022, 95% CI = [0.045, 0.132], *p* < .001).

### Associations of cognitive IIV with cortical thickness IIV

Higher cognitive IIV was associated with higher cortical thickness IIV, reflecting greater heterogeneous cortical thinning across regions (β = 0.036, *SE* = 0.008, 95% CI = [0.020, 0.052], *p* < .001). A significant interaction with clinical severity (β*_INT_* = 0.053, *SE* = 0.017, 95% CI = [0.020, 0.086], *p* = .002; Figure 2C) revealed that the association between higher cognitive IIV and cortical thickness IIV was present in MCI (β = 0.055, *SE* = 0.010, 95% CI = [0.035, 0.075], *p* < .001) but not in CN individuals (β = 0.002, *SE* = 0.014, 95% CI = [-0.024, 0.029], *p* = .87).

### Associations of cognitive IIV with ATN IIV

In the smaller subset of participants with complete ATN biomarker data, higher cognitive IIV was also associated with higher ATN IIV (β = 0.176, *SE* = 0.043, 95% CI = [0.091, 0.262], *p* < .001). Interaction with clinical severity (β*_INT_* = 0.326, *SE* = 0.085, 95% CI = [0.160, 0.493], *p* < .001) showed that a significant effect was observed only in MCI (β = 0.317, *SE* = 0.056, 95% CI = [0.207, 0.428], *p* < .001) but not CN individuals (β = -0.009, *SE* = 0.064, 95% CI = [-0.135, 0.117], *p* = .89, Figure 2D).

## Discussion

In this study, we derived novel measures of AD neuroimaging biomarker IIV and found that higher cognitive IIV was associated with greater biomarker spatial dispersion, with stronger effects in MCI individuals for tau burden, cortical thinning, and combined ATN load. The associations of cognitive IIV with amyloid-β IIV and tau IIV were also amplified in amyloid-β and tau positive individuals, and present even in tau negative individuals. Taken together, these results reflect a close relationship between greater heterogeneity in cognitive performance across domains and greater within-person regional variability in AD-related cerebral pathology, particularly during the phase of prodromal AD with substantial pathological accumulation. Greater cognitive IIV and biomarker IIV likely capture similar disease processes that are clinically meaningful in the heterogeneous pathogenesis of AD.

Extending prior work that has been focused on investigating the relationship between absolute global or regional neuroimaging measures with cognitive IIV [20, 29], our results demonstrated novel links between greater within-person heterogeneity in cognitive performance across domains and dispersion of AD pathology across cortical regions. This is consistent with neuropathological findings that AD-related pathological changes affect specific brain regions in a non-uniform manner [2, 30], with some regions being more vulnerable than others to pathological protein accumulation and distribution throughout the brain [31]. While biomarker thresholds are vital for categorical disease staging, biomarker IIV measures may capture important topographical heterogeneity information. In this context, the extent of heterogeneity in AD pathological distribution that is captured by AD neuroimaging biomarker IIV, may underpin the observed heterogeneity in the clinical manifestation of AD. In particular, cognitive IIV was associated with IIV in all AD biomarkers, suggesting that variation in cognitive performance may reflect broad disease-related disruption spanning both molecular and structural abnormalities that are fundamentally non-uniform long before they become widespread and homogenous.

The strength of these associations differed across biomarkers with stronger effects observed for tau IIV and ATN IIV. This pattern may be understood in light of prior evidence that tau pathology directly correlates with cognitive performance [32, 33], and shows stronger associations with cognitive dysfunction than amyloid-β across the AD continuum [20, 34]. It is also broadly consistent with the amyloid cascade hypothesis, which proposes that the deposition of amyloid may begin earlier in the disease process, often before objective cognitive symptoms, triggering tau pathology and neurodegeneration, leading to cognitive decline and functional impairment [35]. Consistent with this framework, the effects with neuroimaging IIV for all biomarkers, except amyloid-β, were most evident in MCI than CN individuals, indicating a stage-dependent association between regional heterogeneity in tau and neurodegenerative distributions and cognitive performance in AD [35]. Notably, the association between cognitive IIV and tau IIV also remained significant even in tau-negative individuals, suggesting that cognitive IIV is sensitive to subtle variation in tau distribution disease dynamics that global cut-offs do not capture.

More broadly, elevated cognitive IIV, reflecting greater variability in performance across cognitive domains, may be sensitive to the uneven regional dispersion of AD pathology, particularly tau, before widespread cognitive impairment or overt functional deficit. These findings may help explain the heterogenous cognitive profiles commonly observed across the AD continuum. Just as cognitive IIV can be computed from existing neuropsychological batteries without the administration of additional tests, biomarker IIV can also be computed from existing MRI and PET scans without additional neuroimaging scans, thereby enhancing its potential for retrospective analyses in existing research and clinical trials cohorts. The stronger associations observed in MCI and biomarker-positive individuals further suggest that cognitive IIV and biomarker IIV may serve as complementary neuropsychobiological markers to support identification of individuals who may be at a higher risk of progressing to AD dementia.

Several limitations should be acknowledged. Firstly, temporal ordering and potential bidirectionality between cognitive and biomarker IIV could not be established with a cross-sectional analysis. Hence, studies with longitudinal cognitive and neuroimaging data will be needed to determine whether changes in cognitive IIV tend to precede changes in biomarker IIV, or vice versa. Secondly, as analyses were performed in separate biomarker-specific samples of different sizes, comparability of findings across biomarkers may be limited and future studies with more complete biomarker coverage will be needed to confirm the robustness of these findings. Lastly, the present findings derived from ADNI participants may not be generalizable to other populations and would benefit from replication in other independent cohorts.

## Conclusions

Greater heterogeneity in cognitive performance across domains, quantified by elevated cognitive IIV, was associated with greater regional variability in AD-related pathology, particularly tau, with potentially disease stage-dependent effect. These findings provide novel mechanistic insights into the heterogeneous cognitive manifestation in AD beyond global pathological burden and suggest that both cognitive IIV and AD neuroimaging biomarker IIV reflect neuropsychobiological processes implicated in the AD process.

## Abbreviations

A+: Amyloid positive
AD: Alzheimer’s disease
ADNI: Alzheimer’s Disease Neuroimaging Initiative
ATN: Amyloid-Tau-Neurodegeneration
AV-45: Florbetapir
AV-1451: Flortaucipir
CDR: Clinical Dementia Rating
CERAD: Consortium to Establish a Registry for Alzheimer’s Disease
CI: Confidence interval
CN: Cognitively normal
CSF: Cerebrospinal fluid
FDR: False discovery rate
IIV: Intra-individual variability
MCI: Mild cognitive impairment
MMSE: Mini-Mental State Examination
MRI: Magnetic resonance imaging
NFT: Neurofibrillary tangle
PET: Positron emission tomography
RAVLT: Rey Auditory Verbal Learning Test
SE: Standard error
SUV: Standardized uptake value
SUVR: Standardized uptake value ratio
T+: Tau positive
TMT: Trail Making Test

## Declarations

### Ethics approval and consent to participate

Data were obtained from the ADNI, which received ethical approval from institutional review boards at participating sites. All procedures performed in studies involving human participants were in accordance with the ethical standards of the institutional and/or national research committee and with the 1964 Helsinki Declaration and its later amendments or comparable ethical standards.

### Consent for publication

Not applicable.

### Availability of data and materials

The datasets supporting the conclusions of this article are available in the Alzheimer’s Disease Neuroimaging Initiative (ADNI) repository, (https://adni.loni.usc.edu), subject to data use agreement.

### Authors’ contributions

MHT: conceptualisation, data curation, methodology, formal analysis, visualisation, and writing – original draft. MRQT: writing – review and editing. CNK: visualisation and writing – review and editing. CHT: conceptualisation, supervision, funding acquisition, writing – review and editing.

## Acknowledgements

* Data used in preparation of this article were obtained from the Alzheimer’s Disease Neuroimaging Initiative (ADNI) database (adni.loni.usc.edu). As such, the investigators within the ADNI contributed to the design and implementation of ADNI and/or provided data but did not participate in the analysis or writing of this report. A complete listing of ADNI investigators can be found at: https://adni.loni.usc.edu/wp-content/uploads/how_to_apply/ADNI_Acknowledgement_List

